# Surgical Planning and Optimization of Patient-Specific Fontan Grafts With Uncertain Post-Operative Boundary Conditions and Anastomosis Displacement

**DOI:** 10.1101/2021.12.07.21267426

**Authors:** Xiaolong Liu, Narutoshi Hibino, Yue-Hin Loke, Byeol Kim, Paige Mass, Mark D. Fuge, Laura Olivieri, Axel Krieger

## Abstract

**Objective:** Fontan surgical planning involves designing grafts to perform optimized hemodynamic performance for the patient’s long-term health benefit. The uncertainty of post-operative boundary conditions (BC) and graft anastomisis displacements may significantly affect the optimized graft designs and lead to undesired outcomes, especially for hepatic flow distribution (HFD). We aim to develop a computation framework to automatically optimize patient-specific Fontan grafts with the maximized possibility of keeping the post-operative results within clinical acceptable thresholds.

**Methods:** The uncertainties of BC and anastomosis displacements were modeled by using Gaussian distributions according to prior research studies. By parameterizing the Fontan grafts, we built surrogate models of hemodynamic parameters by taking the design parameters and BC as inputs. A two-phased reliability-based robust optimization (RBRO) strategy was developed by combining deterministic optimization (DO) and optimization under uncertainty (OUU) to reduce the computation cost.

**Results:** We evaluated the performance of the RBRO framework by comparing it with the DO method on four Fontan patient cases. The results showed that the surgical plans computed from the proposed method yield up to 79.2% improvement on the reliability of HFD than those from the DO method (*p <* 0.0001). The mean values of iPL and %WSS satisfied the clinically acceptable thresholds.

**Conclusion:** This study demonstrated the effectiveness of our RBRO framework to address uncertainties of BC and anastomosis displacements for Fontan surgical planning.

**Significance:** The technique developed in this paper demonstrates a significant improvement in the reliability of predicted post-operative outcomes for Fontan surgical planning. This planning technique is immediately applicable as a building block to enable technology for optimal long-term outcomes for pediatric Fontan patients and can also be used to other pediatric and adult cardiac surgeries.

## I. Introduction

Fontan surgery is the hallmark operation in the surgical management of single ventricle congenital heart disease patients. In order to establish passive pulmonary blood flow, the surgery involves directing systemic venous blood flow into the pulmonary artery via vascular grafts bypassing the heart [1], [2]. The shape and implantation of Fontan grafts, as shown in Fig. 1A, affect indexed power loss (iPL), hepatic flow distribution (HFD), and wall shear stress (WSS), which correlate to post-surgical complications including decreased exercise capacity [3], pulmonary arteriovenous malformations (PAVM) [4], and thrombosis risk [5], respectively. Tissue-engineered vascular grafts (TEVG) made by three-dimensional (3D) printing and electrospinning nanofibers [6] open the door to enabling personalized vascular grafts that provide optimized hemodynamic performance to reduce the patient’s complications for long-term health benefits. Surgical planning of patient-specific hemodynamically optimized vascular grafts have not yet translated into broad clinical acceptance. In one aspect, the surgical planning processes are primarily operated by engineering teams, and surgical planning has thus been relegated to retrospective post-hoc analysis in a limited number of research centers. The current manual iterative design processes between engineering and clinical teams to optimize patient-specific grafts take a few weeks for the clinical workflow [7], [8]. In another aspect, the accuracy and reliability of predicting post-surgical outcomes require further improvements [9].

**Fig. 1.**
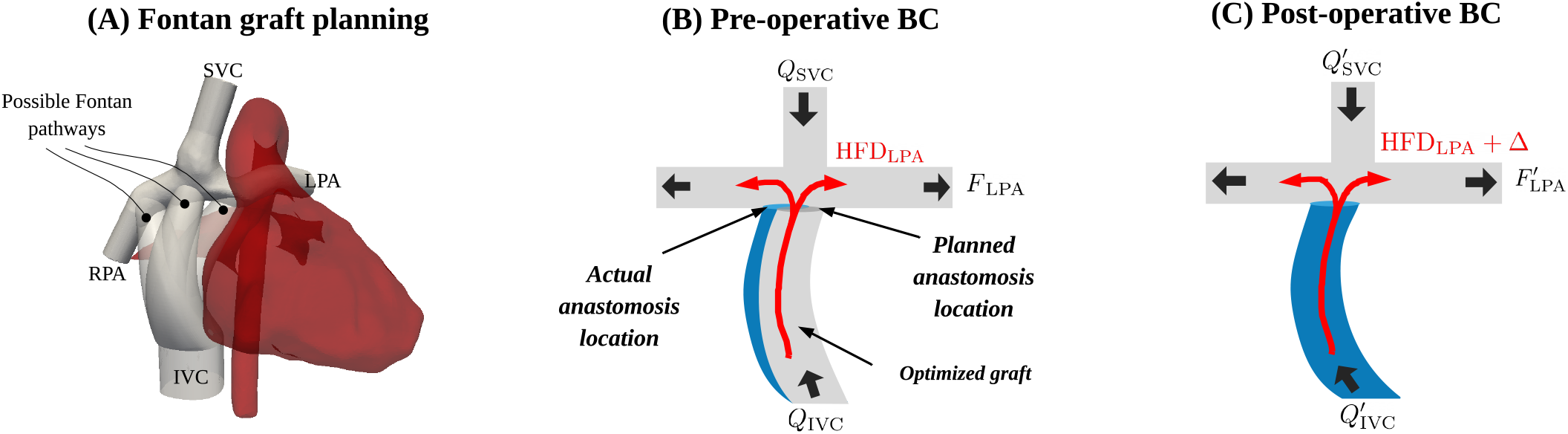
Illustration of Fontan surgical planning and graft implantation under uncertainty. (A) 3D reconstructed patient-specific Fontan model with various possible pathways. Deoxygenated blood flow were directed from the superior vena cava (SVC) and the inferior vena cava (IVC) to the lungs via the left pulmonary artery (LPA) and the right pulmonary artery (RPA). (B) Pre-operative BC, including blood flow rates Q_SVC_, Q_IVC_, and pulmonary artery flow split F_LPA_ were used for setting up the hemodynamic computation. HFD_LPA_ represents the hepatic flow distribution (HFD), which is the percentage of IVC flow to the LPA for the optimized graft at the planned anastomosis location. The blue graft shows the actual ananstomosis location. (C) HFD_LPA_ + Δ. represents the change of HFD due to the uncertainties of the post-operative 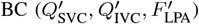 and the anastomosis displacement.

Research efforts have been made to reduce the turnaround time, minimize engineering effort, and improve the hemodynamic performance of the vascular graft surgical planning procedure. Interactive vascular graft modeling tools [10], [11] were developed to substitute general-purpose computer-aided design (CAD) software for speeding up the modeling process of alternative surgical plans. Various computation strategies of Fontan hemodynamics were investigated for improving the computation efficiency while preserving the hemodynamic prediction accuracy [12]. Automatic segmentation methods were developed by taking computed tomography (CT) or magnetic resonance images (MRI) [13] as input to construct 3D cardiovascular structure for reducing manual effort on the image segmentation task.

Instead of manually exploring surgical plans and running post-hoc analysis, automatic Fontan graft optimization techniques were initially developed for idealized models [14], and more recently developed for patient-specific models [15] by parameterizing the graft geometry, building surrogate functions based on the graft parameters to represent high-fidelity Fontan hemodynamics, and searching for the set of graft parameters that can optimize the objective surrogate function. Our prior work [15] has demonstrated that the hemodynamic performance of automatically optimized patient-specific Fontan grafts can match or exceed that of manually optimized grafts, and have a significant reduction of turnaround time (15 hours versus over two weeks). However, current Fontan surgical planning strategies rely on the assumption that the post-operative boundary conditions (BC) are identical to the pre-operative BC, as shown in Fig. 1B. Prior research study [16] has demonstrated that the uncertainty of post-operative BC has a significant impact on HFD of an optimized Fontan graft, as illustrated in Fig. 1C. In addition, even though surgeons can try their best to suture a graft according to the optimized surgical plan, anastomosis displacements are unavoidable, which may also significantly affect HFD.

The objective of this work is to develop a reliable automatic Fontan surgical planning and TEVG shape optimization method, which can tolerate the uncertainties of BC and anastomosis displacements and maximize the possibility of having the hemodynamic parameters of an TEVG implantation within clinically acceptable ranges. Optimization strategies based on the concepts of robustness and reliability have been widely applied in designing engineering systems under uncertainties [17]. Robust optimization (RO) provides solutions presenting less sensitivity to the variability of the design variables [18]. Reliability-based optimization (RBO) computes solutions less prone to failure under the variability constraints of the design parameters [19]. In both approaches, uncertainty quantification (UQ) of the engineering system is performed by sampling-based methods [20], expansion-based methods [21], or most probable point (MPP)-based methods [22]. To reduce the computation cost of UQ process, surrogate modeling methods have been used to substitute high-fidelity simulations [23]. In cardiovascular engineering, the impacts of uncertainties of BC and vessel geometries were investigated for coronary blood flow simulations [24]. To improve the probabilistic distribution modeling of BC, coefficients in parameterized BC models were determined by using clinical data [25]. Different sampling-based strategies were investigated to reduce the computation cost for uncertainty quantification of cardiovascular systems [26], [27]. Despite the advances in UQ for cardiovascular simulation and deterministic optimization (DO) for surgery planning, there is a need of developing a patient-specific cardiovascular graft design that considers various sources of uncertainty and automatically computes optimal surgical plans for surgeons to balance hemodynamic performance, robustness and reliability. In this work, we develop a reliability-based robust optimization (RBRO) framework for patient-specific Fontan surgical planning by leveraging the deterministic TEVG optimization framework developed in our previous work [15]. We introduce uncertain flow split of the left and right pulmonary arteries (LPA, RPA), uncertain flow rate measurements at the inferior vena cava (IVC) and superior vena cava (SVC), and uncertain anastomosis location and orientation of Fontan conduit on the pulmonary artery (PA). We perform constrained optimization to find an optimal set of conduit design parameters that can maximize the probability of keeping HFD within the clinically acceptable thresholds while constraining the mean response values of iPL, WSS within their thresholds.

The main contributions of this work include:

1. We develop the RBRO computation framework of patient-specific Fontan graft planning and optimization under uncertainty for the first time, which moves one step forward towards providing surgeons a reliable tool for pre-operative Fontan surgical planning.
2. We study the effect of warm-starting in the RBRO framework to improve the computation efficiency while preserving the performance of optimal solutions by feeding initial guesses of design parameters, which were computed from the DO solutions.
3. We demonstrate the effectiveness of the RBRO computation framework for Fontan surgical planning on four patient-specific models (n=4) by comparing the performance of Fontan conduits computed from the RBRO method with the performance of conduits obtained from the DO method.
4. We investigate how the RBRO framework works for Fontan patients with highly unbalanced PA flow splits to compute reliable surgical plans. We found that the framework tends to work better for pediatric patients than adult patients.
5. We study how different objective functions in DO affects the warm-start graft designs, and subsequently affects the RBRO results. The objective functions used in DO aim to compute graft designs with balanced HFD or minimized iPL.

## II. Problem Formulation

### A. Research Objective

Given the patient-specific Fontan models, optimization thresholds of hemodynamic parameters, and uncertainty models of BC and anastomosis displacements, our research objective is to automatically design Fontan grafts that maximize the possibility of having all hemodynamic parameters within the clinically acceptable thresholds, which are presented in below.

### B. Hemodynamic Parameters and Optimization Thresholds

The hemodynamic performance of the Fontan pathway is defined by three parameters: 1) HFD; 2) iPL across the Fontan pathway; 3) the percentage of Fontan surface area with non-physiologic wall shear stress, %WSS.

#### 1) Hepatic flow distribution HFD

We use HFD_LPA_ to define HFD. HFD_LPA_ represents the ratio of blood flow from the IVC to the LPA and the total IVC flow.

#### 2) Indexed power loss iPL

iPL is a dimensionless resistive index that correlates with exercise capacity [3]. It is calculated based on the patient’s body surface area (BSA) and the absolute power loss (PL) between the total hemodynamic energy at the inlets (IVC, SVC) and the total hemodynamic energy at the outlets (LPA, RPA):

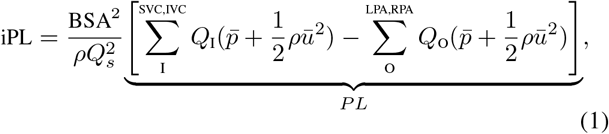

where *Q*_I_ and *Q*_O_ are flow rates at the inlets and outlets respectively, 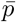 is static pressure, *ρ* is the blood density, *ū* is flow velocity.

#### 3) Percentage of non-physiologic wall shear stress %WSS

Oversized Fontan conduits can lead to low WSS that correlates with neoinitimal hyperplasia and thrombosis [28]. To prevent conduit oversizing, %WSS was introduced to measure the percentage of low WSS area on the luminal surface of Fontan conduits [8]:

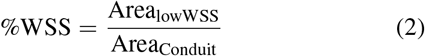

#### 4) Thresholds for Fontan Graft Optimization

- HFD_balanced_: 40% < HFD_LPA_ < 60%. The ideal HFD with 50% IVC flow to LPA is not always feasible. Based on Haggerty et al.’s computational fluid dynamics (CFD) study of hemodynamic parameters in 100 Fontan patients [29], the mean LPA split is 44% with interquartile range 31% to 57%. We aimed to have an acceptable HFD range to match this cohort as 40% ∼ 60%. In the following text, we use HFD_balanced_ to represent 40% < HFD_LPA_ < 60%.
- iPL < 0.03. Based on [29], the mean iPL was 0.037, and the median iPL was 0.031. We set the iPL threshold at 0.03.
- %WSS < 10%. The normal physiologic range of WSS for venous flow is 1 ∼ 10 dynes/cm^2^ (0.1 ∼ 1 Pa) [30]. Area_lowWSS_ in (2) represents the surface areas with WSS below 1 dynes/cm^2.^

### C. 3D Fontan Modeling and CFD Simulation

This study was approved by the Institutional Review Boards (IRBs) of all institutions with the IRB Protocol Number Pro00013357. Cardiovascular magnetic resonance imaging (CMR) datasets from four patients who had undergone Fontan operation were anonymized and exported. By using angiography data with late-phase, non-gated, breath-held acquisition with pixel size 1.4 ×1.4 mm, 3D anatomic replicas of the Fontan and proximal thoracic vasculature were created, as illustrated in Fig. 2. Phase contrast images were used to extract flow curves for the inlet and outlet BC for each patient.

**Fig. 2.**
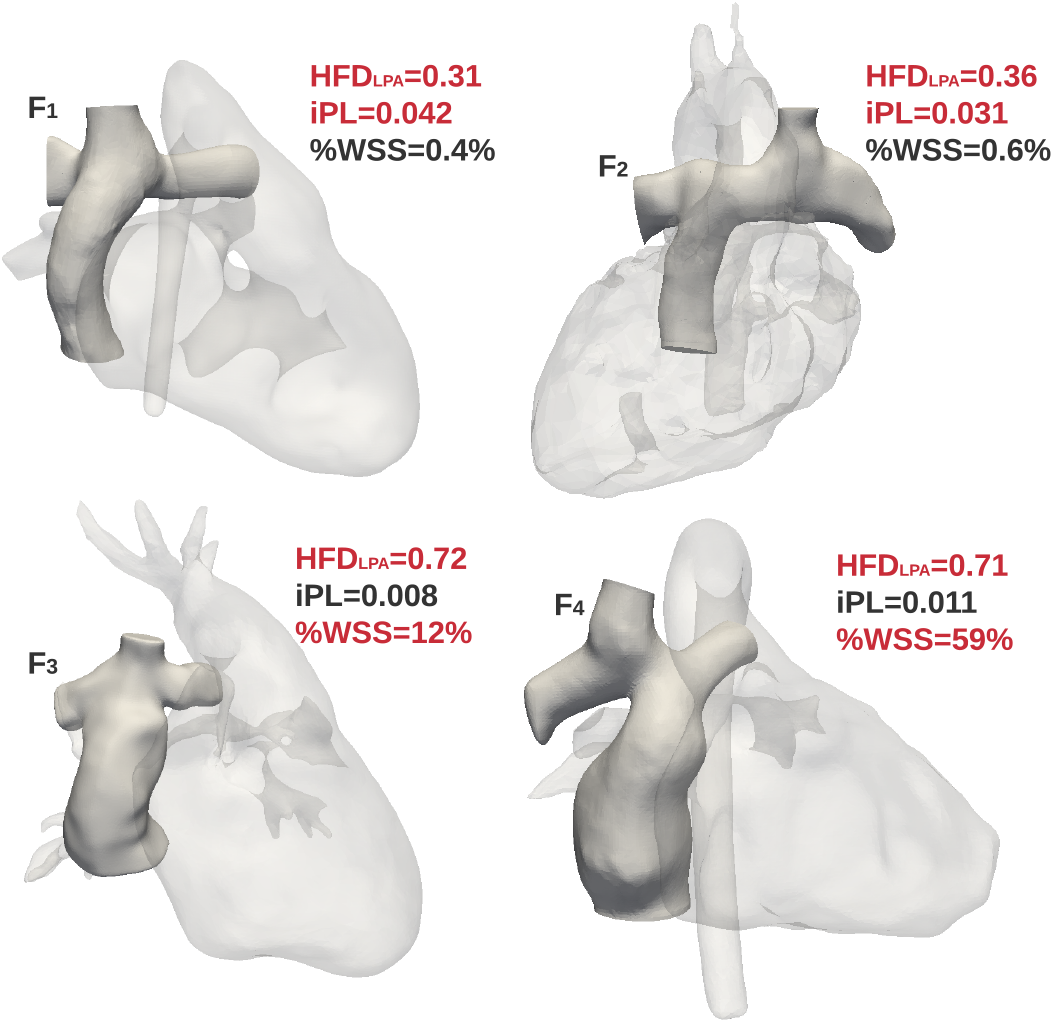
Three-dimensional representation and hemodynamic results of original Fontans. A cohort of Fontan patients (n=4) were retrospectively collected and digitally processed into 3D models for CFD simulation. The cohort consisted of 2 extracardiac-type Fontans (F_1_, F_2_), and 2 lateral tunnel-type Fontans (F_3_, F_4_). The original Fontan conduits were removed from the model for the surgical planning task. The highlighted hemodynamic parameters in red color were considered outside the thresholds.

CFD simulation methodology is as detailed in our prior study [15]. We employed the open source CFD software OpenFOAM [31] to compute the Fontan hemodynamics. Time-averaged IVC and SVC flow rates *Q*_IVC_, *Q*_SVC_ were derived from the extracted flow curves and prescribed at the inlets. Time-averaged LPA and RPA flow rates were calculated by using the PA flow split *F*_LPA_ = *Q*_LPA_/*Q*_Total_ with total venous flow

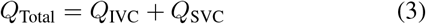

and prescribed at the outlets, as shown in Fig. 1B. Each patient’s BC are listed in the supplementary material. Massless infinitesimal particles were released at IVC to trace the hepatic blood flow. The HFD can be represented by the ratio of particle numbers that arrive at LPA (N_LPA_) and RPA (N_RPA_):

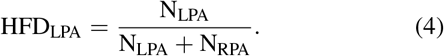

iPL and %WSS were calculated based on (1), (2), and the solved computation domain. The parameters highlighted in red color in Fig. 2 were considered beyond the threshold.

### D. Uncertainty Modeling

For surgical planning, the original Fontan conduits shown in Fig. 2 were removed from the models to create a superior cavopulmonary connection (SCPC) model. The studies in [9], [32] demonstrate that the change in post-operative BC significantly affects the hemodynamic results, especially for HFD. Approximate ± 20% differences between pre- and post-operative BC were demonstrated in a short-term study [9]. In a long-term study [32], the changes in BC were much larger due to the patient’s growth. In this study, we only focus on the uncertainty of post-operative BC in the short term. We assume the post-operative BC follow the Gaussian distribution with the pre-operative BC values as mean and the standard deviation 3σ_BC_ = 20%. The cardiac output *Q*_Total_ is constant.

Based on the pre-operative BC and the uncertainty model, the post-operative BC are

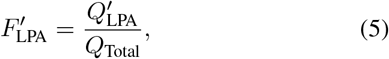

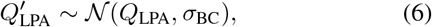

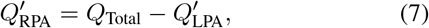

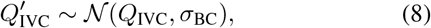

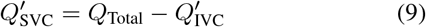

In addition to the BC uncertainty, anastomosis displacements of Fontan conduits also degrade the hemodynamic performance of an optimized Fontan pathway. Fig. 3A and Fig. 3B illustrate two types of displacements, i.e., translational displacement *δ*_*d*_ and rotational displacement *δ*_*ϕ*_, respectively. The grafts in gray color represent the prescribed surgical plan, while the blue ones demonstrate the actual surgical implantation. The graft implementation accuracy relies on the surgeon’s skill and assistive tools, such as paper rulers. Clinical data about surgical implantation accuracy are currently unavailable. Based on the suggestion from our medical co-authors, we assume that the maximum translational displacement is 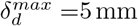 and the maximum rotational displacement is 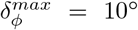. We use Gaussian distribution to model the anastomosis uncertainty:

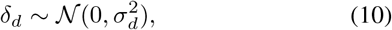

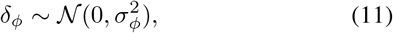

where 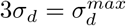 and 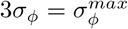.

**Fig. 3.**
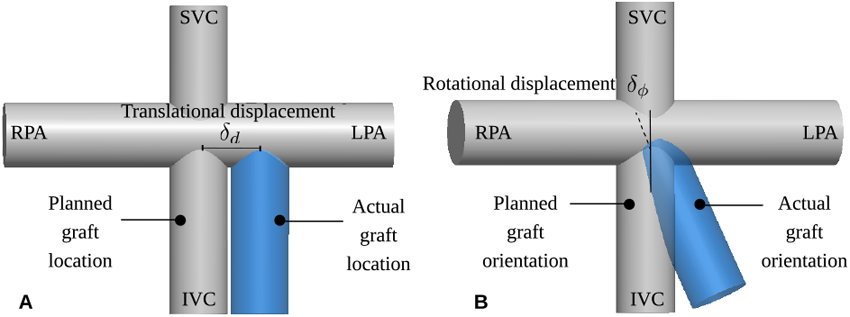
Simplified Fontan models to illustrate the translational and rotational anastomosis displacements. (A) *δ*_*d*_ represents the uncertainty of translational displacement. (B) *δ*_*ϕ*_ represents the uncertainty of rotational displacement.

## III. Reliability-based Robust Optimization Framework

### A. Overview of Computation Framework

The RBRO framework consists of four main components, as shown in Fig. 4: (A) framework input; (B) Fontan pathway parameterization; (C) surrogate model generation for hemodynamic parameters; and (D) optimization of Fontan pathways. 3D patient-specific models were pre-processed to remove the existing Fontan pathways for new graft planning. The pre-operative BC include the percentage of outflow to the LPA *F*_LPA_, inlet flow rates *Q*_IVC_ and *Q*_SVC_, and the estimated post-operative BC 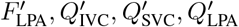 and 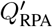. The Fontan pathway parameterization defines the design space of grafts to explore various surgical plans. Computing the hemodynamic parameters directly from high-fidelity simulations is computationally expensive, which makes the optimization process formidable. Surrogate models of high-fidelity hemodynamic simulation significantly reduce the computation time while preserving the prediction accuracy. By taking the graft design space, surrogate models and the uncertainty models as the inputs, we developed a two-step optimization strategy, including a DO step and an OUU step, to explore the reliability and robustness of the Fontan graft designs.

**Fig. 4.**
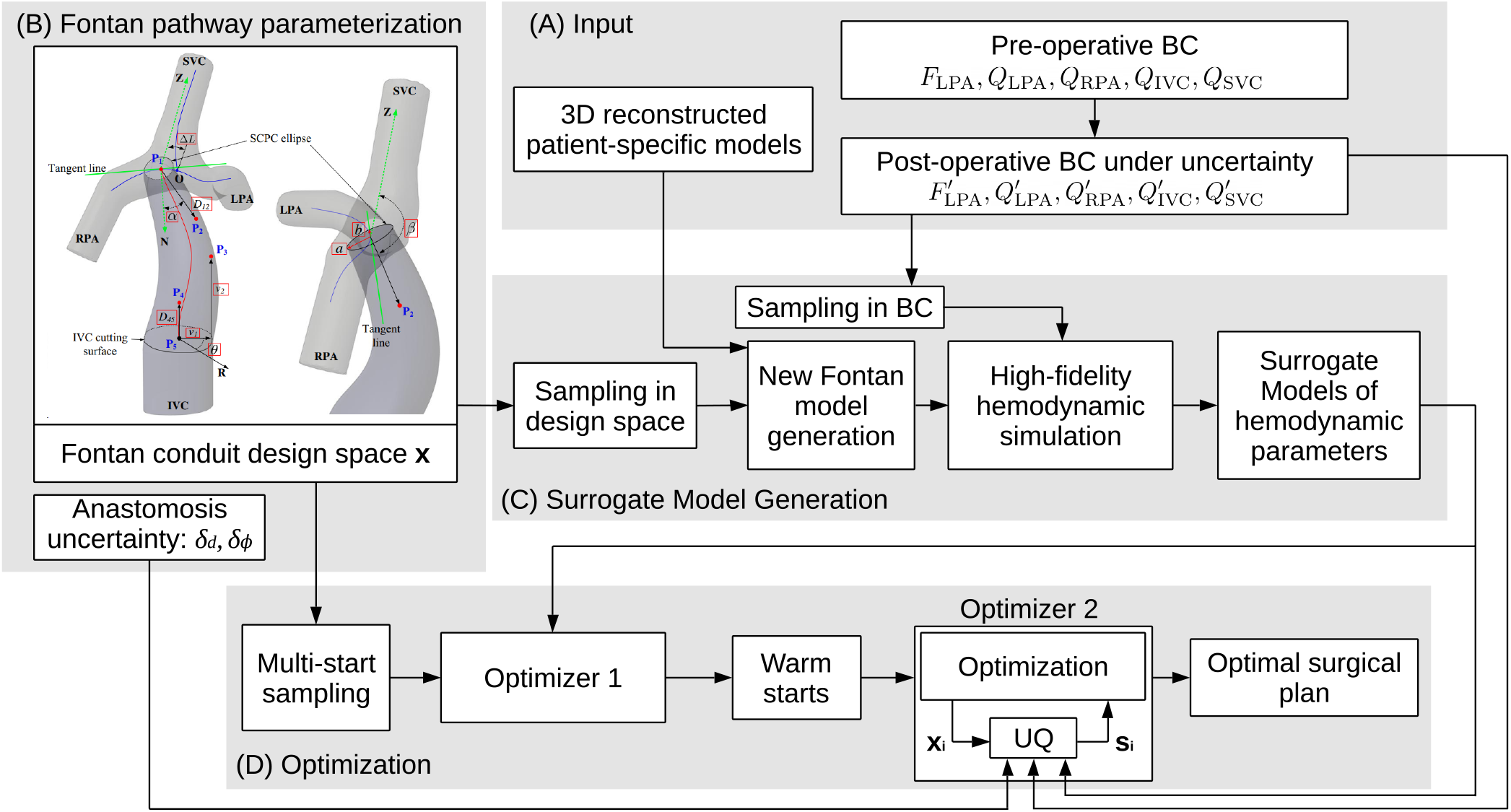
Schematic of the RBRO computation framework. (A) The inputs of the framework include 3D Fontan models, the pre-operative BC, and the uncertainty models of BC. (B) Fontan pathway parameterization creates the design space **x** for automatically exploring the conduit geometry, anastomosis location and orientation. The uncertainty models of anastomosis can also be considered as the inputs of the framework. (C) The surrogate model generation involves sampling in the design space and the space of uncertain parameters, computing the hemodynamic results, and applying Gaussian process regression to learn the hemodynamic responses of different inputs. (D) The RBRO optimization process includes two optimizers. The optimizer 1 performs DO to generate warm starts for the optimizer 2. The optimizer 2 performs OUU to compute the final optimal surgical plans.

### B. Fontan Conduit Parameterization

We created a 10-dimensional design space to parameterize a patient-specific Fontan conduit in our prior study [15] as shown in Fig. 4B. The design parameters **x** = {*a, b, α, β*, Δ*L, D*_12_, *D*_45_, *v*_1_, *v*_2_, *θ*} ∈ 𝒟_***x***_ are highlighted in the red boxes. *a* and *b* define the size of the conduit on the side of the superior cavopulmonary connection (SCPC). *α* and *β* define the conduit’s anastomosis angle. Δ*L* represents the anastomosis location on the PA. The conduit pathway is defined by a fourth-order Bézier curve, whose shape is controlled by spatial points **P**_1_(Δ*L*), **P**_2_(Δ*L, α, β, D*_12_), **P**_3_(*v*_1_, *v*_2_, *θ*), **P**_4_(*D*_45_), **P**_5_. The conduit 3D reconstruction is detailed in [15].

Three parameters Δ*L, α, β* in the design space are subjected to uncertainty. According to (10) and (11), the probability distributions for each set of sampled parameters Δ*L*_*s*_, *α*_*s*_, *β*_*s*_ are represented by

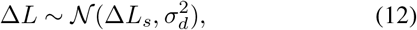

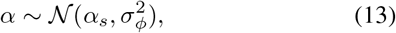

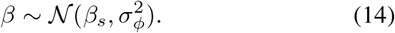

### C. Surrogate Model Generation

To reduce the computation cost for evaluating hemodynamic performance and graft implantation feasibility for various conduit designs, Gaussian process regression with radial-basis functions (RBFs) was used to generate surrogate models for HFD, iPL, %WSS as well as the geometrical interference between the conduit and the heart model (InDep) and the conduit model quality (Nv) [15]. Maximum Likelihood Estimation (MLE) is used to find the optimal values of the hyper-parameters of RBFs governing the trend and correlation functions. In addition to the 10 design parameters **x**, two uncertainty variables of BC *F*_LPA_ and *Q*_IVC_ are added as input arguments of the surrogate models. By representing

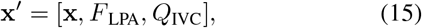

the surrogate models are formulated as

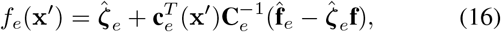

where *e* = {iPL, HFD_LPA_, %WSS, N_v_, InDep}, **C**_*e*_ is the covariance matrix, **c**_*e*_(**x**^′^) is covarance vector, 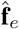 is the vector of training data. **f** is a unity vector. 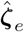 is the generalized least squares estimate of the mean response [33].

Latin hypercube sampling (LHS) was employed to sample **x**^′^. We have investigated the surrogate model accuracy based on the 10-dimentional design space in our prior study [15]. The results indicated that 2000 training samples can provide a good trade-off between surrogate prediction accuracy and the data scale. Although two additional parameters are added in this study, the cross-validation results for the surrogate models with 12 input parameters demonstrated similar accuracy to the results for the surrogate models with 10 design parameters. Therefore, in this study, 2000∼3000 samples that require running high-fidelity hemodynamic simulations for each sample were used to train the surrogate models for each patient.

### D. Optimization Under Uncertainty

We aim to compute Fontan conduits that can resist the influence of the uncertainties of BC and anastomosis displacements to satisfy the thresholds of Fontan hemodynamics presented in Section II-B4. Because HFD is the most sensitive Fontan hemodynamic parameter under the presence of uncertainties, the objective of the Fontan conduit optimization is to search for conduit design parameters that maximize the probability of HFD_balanced_.

#### 1) Optimization Strategy

To search for globally optimal solutions under uncertainty, an intuitive optimization strategy is to first sample a sufficient number (*N* = 600 [15]) of initial guesses distributed in the design space. Starting from each initial guess **x**_0_ as shown in Fig. 5, an optimizer then searches in different directions and generates a new **x**_*k*_ at the *k*^*th*^ iteration with the maximum iteration number *K*. This UQ process generates *M* samples in the uncertainty space, which when combined with the design parameters **x**_*k*_, can compute the statistics of the response functions **S**_*r*._ Assuming *M* = 100, *K* = 200, the total number of simulations is approximately *N* × *M* × *K*=12, 000, 000. Our high performance computing cluster is unable to handle that many files for each run. To improve the computation efficiency, an alternative optimization strategy is to employ DO to compute optimal solutions as the warm starts for OUU. An estimated number of the total simulations is *N* × *K* + 10 × *K* × *M* = 320, 000 which is 37.5 times more efficient than the first strategy. Fig. 4D shows the optimization workflow. The first optimizer solves the DO problem, and the second optimizer solves the OUU problem with warm starts from DO.

**Fig. 5.**
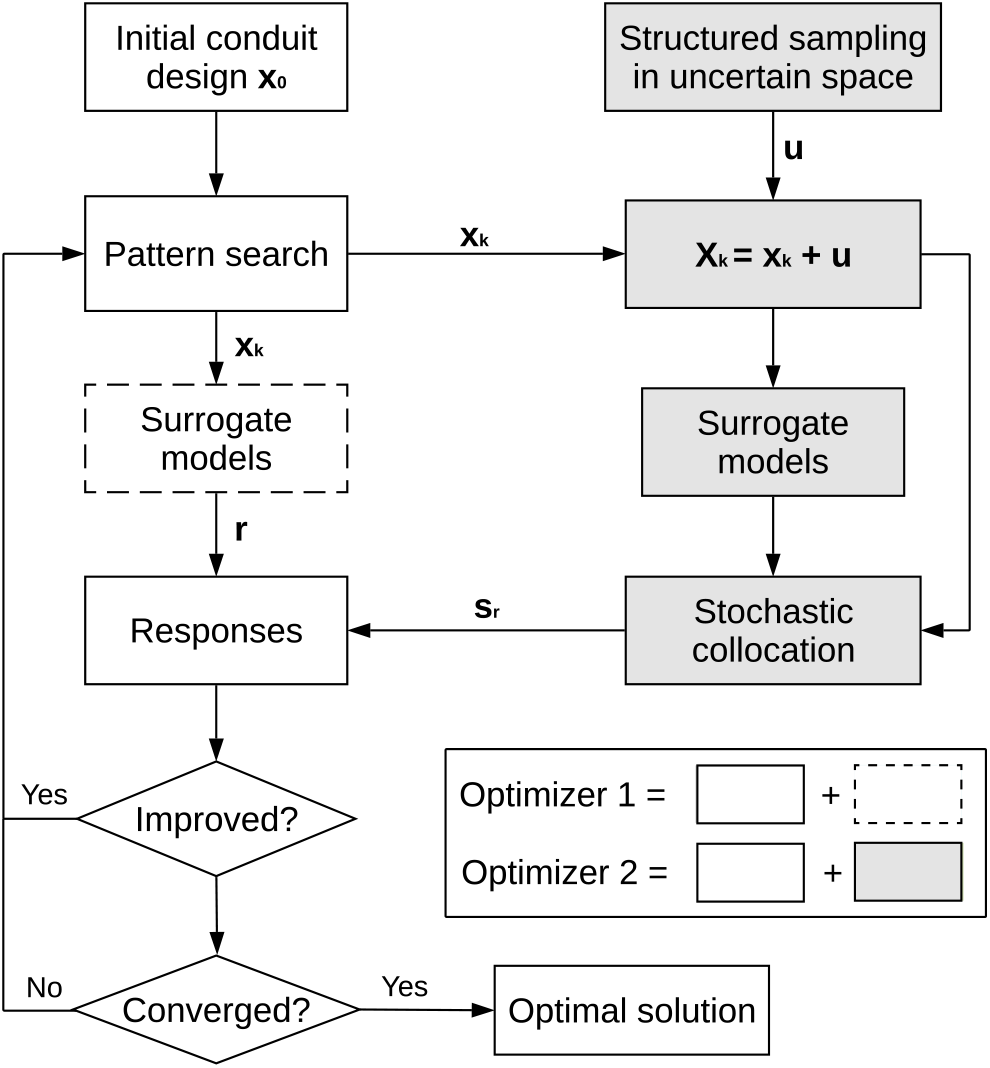
Block diagram of the optimizer 1 and the optimizer 2. The optimizer 1 represents the DO process, which does not consider the uncertainty of BC and anastomosis. The optimizer 2 performs uncertainty quantification (UQ) with the uncertainty space **u** on each set of explored design parameters **x**_*k*_ to generate statistic responses of the hemodynamic parameters.

#### 2) Deterministic Optimization

The optimizer 1 shown in Fig. 4D and detailed in Fig. 5 performs DO on a set of initial conduit designs (*N* = 600) generated by the LHS method.

The DO problem is formulated as:

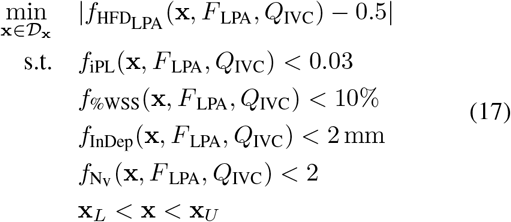

where the objective function aims to find conduit designs with HFD_LPA_ = 0.5, *F*_LPA_ and *Q*_IVC_ are with deterministic values, the surrogate functions 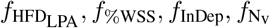 are defined in (16), the thresholds in the constraints are defined in Section II-B4, **x**_*L*_ and **x**_*U*_ are the lower and upper bounds of **x**. We employed the asynchronous parallel pattern search (APPS) method [34] to generate new search points **x**_*k*_, as shown in Fig. 5, in the design space for finding the optimal solutions.

#### 3) Optimization Under Uncertainty

The best 10 solutions from DO are used as the warms starts for the optimization formulated as:

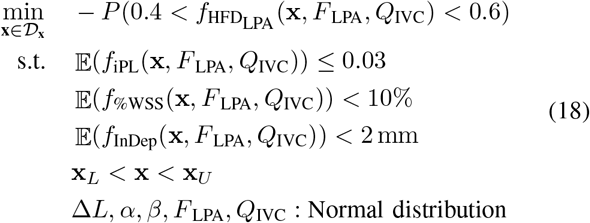

We converted the maximization problem to a minimization problem by adding a minus sign to 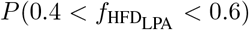. 𝔼(·) represents the expectation operator.

In each optimization iteration in the optimizer 2 shown in Fig. 5, the uncertainty space

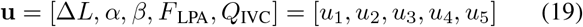

is sampled to evaluate the statistics at **x**_*k*_. We employ the stochastic collocation method for uncertainty quantification because of its higher efficiency and faster convergence rate than that of sampling-based methods [21].

The stochastic collocation (SC) is represented by Lagrange interpolation functions with known coefficients from the sampled uncertain variables ***u*** = [**u**_1_, **u**_2_, …, **u**_*n*_] (n represents the sample number) and their corresponding response vector **r**. We used the Smolyak-type sparse grid method with the grid level at 2 for generating the samples [35]. Defining *r*_*i*_ as the *i*^*th*^ element in **r** that represents the response values at interpolation points, the SC is formulated as

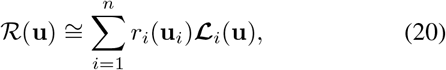

where **ℒ**_*i*_(**u**) is the *i*^*th*^ Lagrange polynomial:

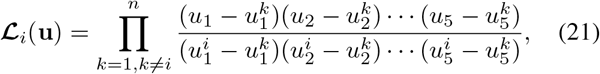

the superscripts of u represent the sample index, and **ℒ**_*i*_(**u**_*j*_) = *δ*_*i,j*_, and *δ*_*i,j*_ is the Kronecker Delta.

The moments of (20) can be derived in closed form:

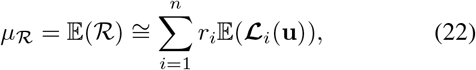

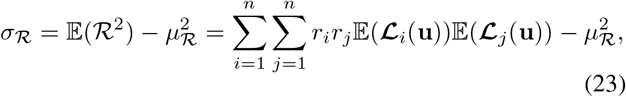

To evaluate the probabilities, 10^5^ samples were applied to the stochastic expansion in (20). The RBRO framework was implemented based on the Dakota software package [36].

## IV. Results

### A. Comparison of Results from DO and OUU

We first examine the performance of Fontan conduit designs computed from DO, and compare them with the performance of the conduit designs from OUU to evaluate the effectiveness of the RBRO framework. Fig. 6A-D show reliability of HFD *P* (HFD_balanced_) of the optimized conduit designs by using UQ for the four patient cases. The 10 best designs from DO as the warm starts were computed for OUU by ranking |HFD_LPA_ - 0.5| of each optimized conduit design. The design orders were descendingly sorted according to *P*(HFD_balanced_). The original order indices of DO design are labeled on the top of each bar group. The gray and red bars represent the results from DO and OUU, respectively. The result demonstrates the original rankings of DO designs are unable to guarantee their reliability. For certain patients, such as F_4_, the best original DO design has the worst reliability. The RBRO framework can improve the reliability of DO solutions up to 56.7%, 9.3%, 6.6%, and 79.2% for the four patients, respectively.

**Fig. 6.**
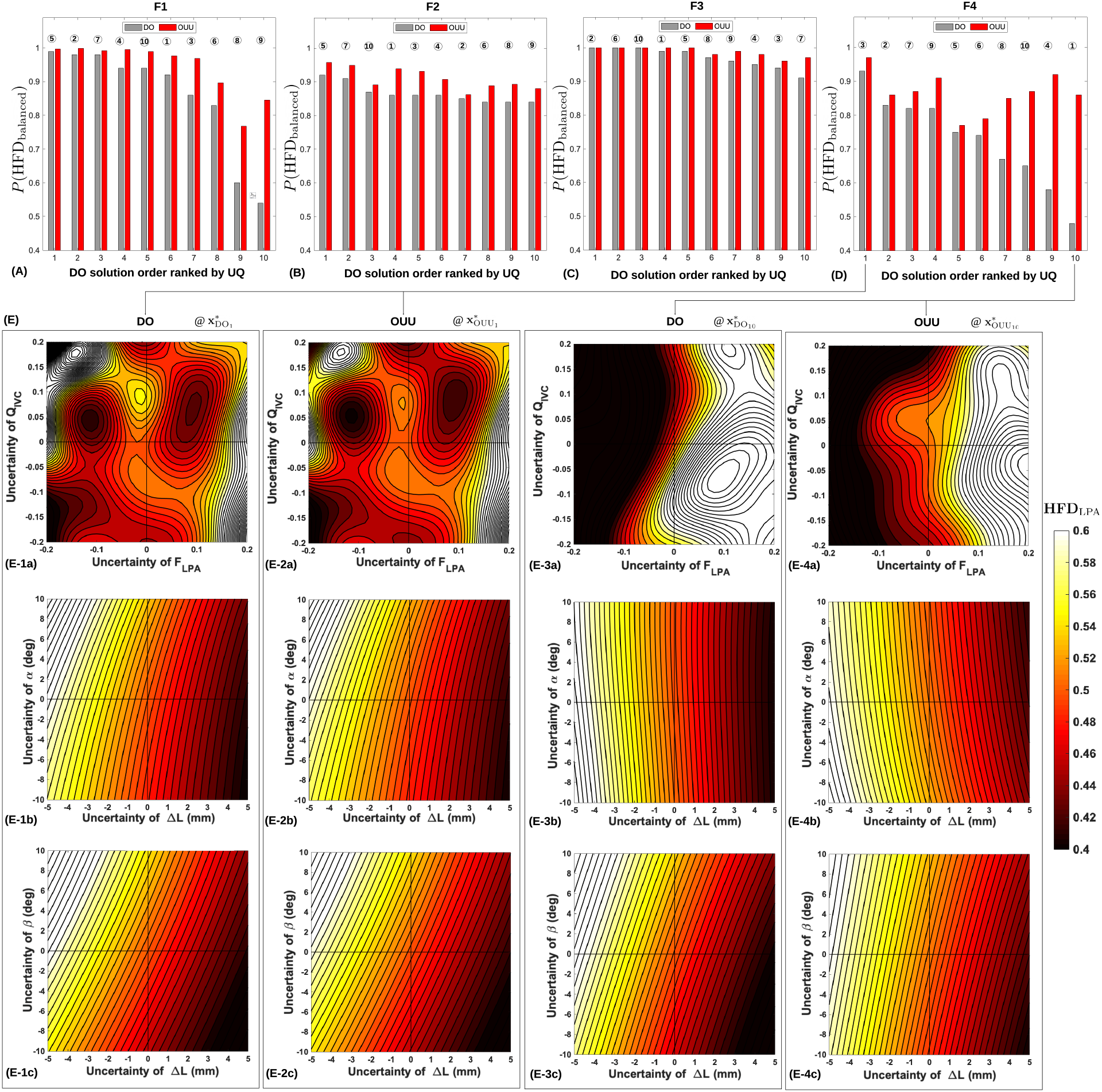
The probability of having balanced HFD P(HFD_balanced_) of optimized graft designs by using deterministic optimization (DO) and optimization under uncertainty (OUU) methods for each patient. The result for the patient F_1_ is shown in (A). The y-axis represents the probability of HFD_LPA_ within the thresholds. The x-axis represents DO design order ranking from the most reliable HFD to the least reliable HFD by using UQ. The 10 DO designs are the warm starts for OUU. The circled numbers above the bars represent the original ranking of the DO designs according to the objective function of (17). The same analysis for the patients F_2_, F_3_ and F_4_ are shown in (B), (C), and (D), respectively. (E) illustrates how the uncertain parameters **u** affect HFD_LPA_ for the optimized designs in the groups #1 and #10 of F_4_. The three rows show HFD_LPA_(*F*_LPA_, *Q*_IVC_, **x**), HFD_LPA_(Δ*L, α*, **x**), HFD_LPA_(Δ*L, β*, **x**). **x** represents the graft design parameters 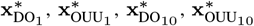 in the four columns, respectively.

Fig. 6E demonstrates how HFD_LPA_ changes under the uncertain parameters **u** at 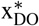 and 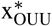, which are the optimized design parameters from DO and OUU, respectively. In this example, we compared four conduit designs from the groups #1 (top-ranked DO design) and #10 (best original DO design) in F_4_. In each column of Fig. 6E, the three rows from top to bottom represent functions of HFD_LPA_(*F*_LPA_, *Q*_IVC_, **x**), HFD_LPA_(Δ*L, α*, **x**), and HFD_LPA_(Δ*L, β*, **x**). In the contour map, the white color indicates HFD_LPA_ close to or higher than 0.6, and the black color indicates HFD_LPA_ close to or lower than 0.4. The OUU solutions 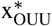 exhibits significantly wider range of HFD_LPA_ within the thresholds than 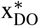, especially for the group #10, by comparing Fig. 6E-3a and Fig. 6E-4a, Fig. 6E-3b and Fig. 6E-4b, and Fig. 6E-3c and Fig. 6E-4c. The DO solution in the group #1 already provided high reliability of HFD, OUU demonstrates minor improvement on the reliability against *α, β*, Δ*L*, as shown in Fig. 6E-1b, Fig. 6E-2b, Fig. 6E-1c, and Fig. 6E-2c. However, a small improvement on the handling uncertainty of BC can be observed in Fig. 6E-1a and Fig. 6E-2a.

The top-ranked conduit designs from DO and OUU as well as the best original DO designs for the four patients are presented in Table I. The top-ranked design parameters of DO and OUU for each patient are close to small adjustments in conduit size (F_1_, F_2_, F_4_), anastomosis angle (F_2_, F_4_), anastomosis location (F_1_). The best original DO designs have lower HFD reliability and higher iPL than the top-ranked DO designs. Fig. 7 shows the top-ranked DO and OUU conduit designs, which are represented in gray and red colors, respectively. The similar design parameters lead to comparable hemodynamic performance with 0%∼4% improvements on *P*(HFD_balanced_), 0%∼9.4% improvements on 𝔼(iPL) by using RBRO framework.

**TABLE I.** Design parameters and Fontan hemodynamic responses of optimal design. Top-ranked DO design, top-ranked OUU design, and the best original DO design for each patient are presented.

| Optimal designs | Design parameters |  |  |  |  |  |  |  |  |  | Responses |  |  |
| --- | --- | --- | --- | --- | --- | --- | --- | --- | --- | --- | --- | --- | --- |
| | a (mm) | b (mm) | $\alpha$ (rad) | $\beta$ (rad) | $\Delta L$ | $D_{12}$ | $D_{45}$ | $v_1$ (mm) | $v_2$ (mm) | $\theta$ (rad) | $P(\text{HFD}_{\text{balanced}})$ | $\mathbb{E}(\text{iPL})$ | $\mathbb{E}(\% \text{WSS})$ (%) |
| F<sub>1</sub>, top-ranked DO | 12.29 | 10.44 | 1 | -0.66 | 85.54 | 1 | 1 | 40.74 | 14.67 | 3.37 | 0.99 | 0.026 | 0.97 |
| F<sub>1</sub>, top-ranked OOU | 11.44 | 10.02 | 1 | -0.66 | 84.50 | 1 | 0.75 | 40.74 | 14.67 | 3.37 | 1 | 0.023 | 0.98 |
| F<sub>1</sub>, best original DO | 10.23 | 7.77 | 0.45 | -0.52 | 86.32 | 0.30 | 0.92 | 21.79 | 38.05 | 3.83 | 0.92 | 0.028 | 1.62 |
| F<sub>2</sub>, top-ranked DO | 7.35 | 9.57 | 0.76 | -0.03 | 186.36 | 0.36 | 0.98 | 39.62 | 17.57 | 5.15 | 0.92 | 0.022 | 3.05 |
| F<sub>2</sub>, top-ranked OOU | 8.49 | 10 | 0.01 | 0.53 | 186.36 | 0.30 | 0.85 | 39.62 | 17.27 | 5.35 | 0.96 | 0.019 | 2.83 |
| F<sub>2</sub>, best original DO | 9.84 | 7.57 | 1 | -0.38 | 166.54 | 1 | 0.25 | 38.82 | 11.60 | 1.02 | 0.86 | 0.017 | 2.63 |
| F<sub>3</sub>, top-ranked DO | 8.61 | 10 | 1 | -0.06 | 73.11 | 1 | 0.48 | 37.99 | 35.89 | 4.36 | 1 | 0.014 | 0.91 |
| F<sub>3</sub>, top-ranked OOU | 8.61 | 10 | 1 | -0.06 | 73.11 | 1 | 0.48 | 37.99 | 35.89 | 4.36 | 1 | 0.014 | 0.91 |
| F<sub>3</sub>, best original DO | 7.82 | 6.99 | 0.64 | -0.26 | 58.21 | 0.48 | 0.21 | 44.60 | 29.38 | 4.07 | 0.99 | 0.024 | 0.76 |
| F<sub>4</sub>, top-ranked DO | 8.32 | 9.65 | 0.96 | 0.03 | 146.37 | 0.57 | 0.47 | 46.75 | 22.85 | 3.28 | 0.93 | 0.015 | 3.5 |
| F<sub>4</sub>, top-ranked OOU | 8.32 | 10.65 | 1 | 0.03 | 146.37 | 0.57 | 0.47 | 46.75 | 22.85 | 3.28 | 0.97 | 0.014 | 3.8 |
| F<sub>4</sub>, best original DO | 8.85 | 9.11 | 0.79 | -0.93 | 127.01 | 0.1 | 1 | 8.89 | 21.99 | 0.96 | 0.48 | 0.018 | 5.5 |

**Fig. 7.**
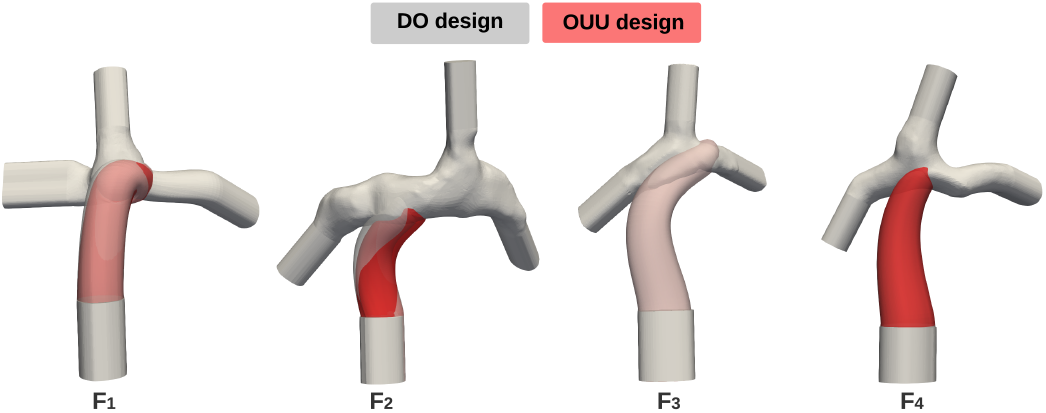
Illustration of the top-ranked DO and OUU designs for the 4 patients. It is hard to tell the geometrical differences between DO and OUU designs for F_3_ and F_4_, because the design parameters are identical for DO and OUU in F_3_ (see Table I) and the graft diameter of the OUU design in F_4_ is slightly larger than that of the DO design with all the other design parameters almost the same.

### B. OUU for Patients with Highly Unbalanced F_LPA_

It is not always feasible to design Fontan conduits with HFD_LPA_ = 0.5 if the patient has highly unbalanced PA split *F*_LPA_. We are interested in investigating the performance of RBRO framework for applying to these extreme cases.

HFD_LPA can_ be formulated by:

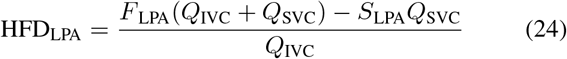

where *F*_LPA_(*Q*_IVC_ + *Q*_SVC_) represents the venous flow rate to the LPA, *S*_LPA_*Q*_SVC_ represents the flow rate from SVC to LPA. The best HFD_LPA_ can be found by varying the fraction *S*_LPA_ from 0 to 1 [37].

Fig. 8 visualizes the best possible HFD_LPA_ by giving specific *F*_LPA_ and the percentage of IVC flow in the total systemic venous flow *Q*_IVC_/(*Q*_IVC_ + *Q*_SVC_) based on (24). We used 0.5∼0.8 as the range of *Q*_IVC_/(*Q*_IVC_ + *Q*_SVC_) to represent the spectrum from pediatric patients to adult patients. The red, white, and blue regions indicate that the best HFD_LPA_ is higher than 0.5, equal to 0.5, and lower than 0.5, respectively. We marked the locations of the four patient cases in Fig. 8 according to their BC. It shows that the best theoretical HFD_LPA_ for F_1_, F_3_, and F_4_ are 0.5. Although F_2_ is at the boundary line with the value slightly higher than 0.5, we were able to compute conduit designs with HFD_LPA_ = 0.5 most likely because of the imperfection of the Lagrangian particle tracking algorithm for the HFD computation. To prepare the extreme cases with highly unbalanced *F*_LPA_, we employed the Fontan geometry of F_1_ and changed its original BC to the values indicated in T_1_ (*F*_LPA_ = 0.74, *Q*_IVC_/(*Q*_IVC_ + *Q*_SVC_)= 0.72) and T_2_ (*F*_LPA_ = 0.2, *Q*_IVC_/(*Q*_IVC_ + *Q*_SVC_) = 0.54), as illustrated in Fig. 8.

**Fig. 8.**
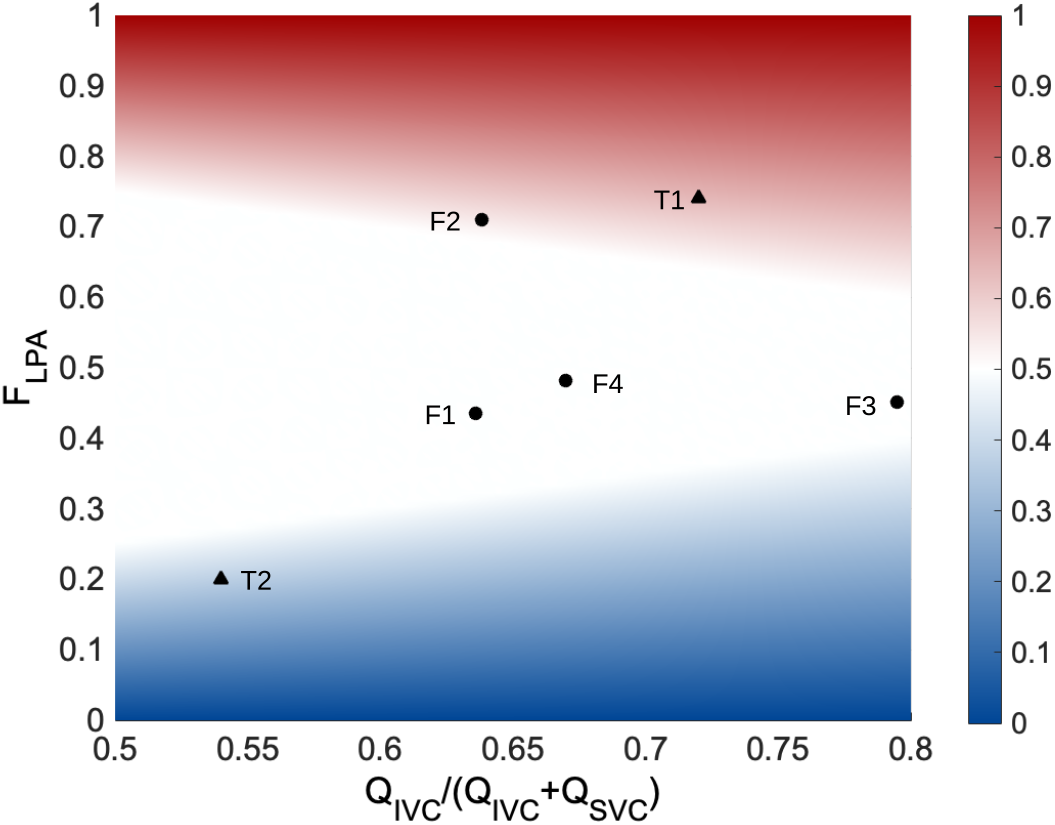
The feasibility of designing grafts with HFD_LPA_ = 50% under different patients’ blood flow conditions. The x-axis represents the percentage of Q_IVC_ in the total systemic venous flow (Q_IVC_ +Q_SVC_). The range 0.5∼0.8 represents the patient spectrum from pediatric to adult. The y-axis represents F_LPA_. The HFD_LPA_ map is calculated by using (24). The locations of F_1_, F_2_, F_3_, and F_4_ correspond with their pre-operative BC. T_1_ and T_2_ represent two extreme cases with highly unbalanced PA flow splits.

Fig. 9 shows DO (black dots) and OUU (red dots) results for T_1_ and T_2_. The dash lines indicate the DO solutions that the OUU solutions were computed from. As shown in Fig. 9A, OUU can hardly improve the conduit designs based on the DO solutions for T_1_ even though the *P* (HFD_balanced_) values are below 0.7. But for T_2_, OUU significantly improves the reliability of the conduit designs by comparing with the DO results. We also noticed that all the conduit designs yields much higher power loss (exceeded the 0.03 iPL threshold) in T_2_. There is minor concern about %WSS since all values are far below the threshold 0.1.

**Fig. 9.**
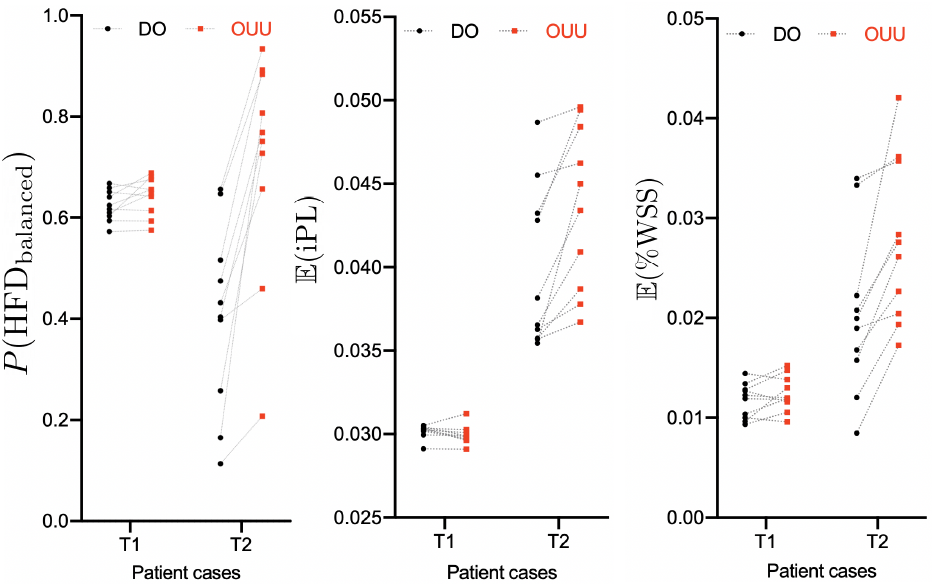
Fontan graft optimization results for the patient cases T_1_ and T_2_ with highly unbalanced pulmonary artery flow splits. Each black dot represents an optimized graft design that was computed from DO. Each red dot represents an optimized graft design that was computed from OUU with a DO solution as a warm start. The dash lines indicate that specific DO designs were used for OUU computation.

### C. OUU with Different Objective Function in DO

We formulated the optimizer 1 by optimizing HFD in the objective function (17) for the RBRO framework. We are interested in studying the influence of using a different DO formulation, i.e., minimizing iPL as the objective function:

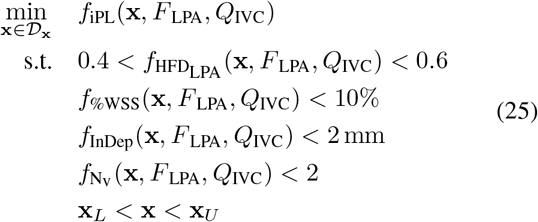

Instead of pursuing a perfectly balanced HFD in DO, (25) minimizes iPL while constraining HFD within the threshold.

Fig. 10 demonstrates how the changing of objective function in DO affect DO and OUU results. Fig. 10A and Fig. 10D compare the reliability of HFD and robustness of iPL in DO designs with min |HFD_LPA_ − 0.5| (red bars) and min iPL (gray bars) as the objective functions. We represent mean and standard deviation of each data group as mean ± standard deviation in the following text. The unpaired 2-tailed t-tests were used to compare the results between the red and gray groups. A p-value < .05 was considered statistically significant. In Fig. 10A, we found except for the patient F_1_ (red: 0.86 ±0.03, gray: 0.80 ±0.03, *p* = 0.49), the red groups statistically perform better than the gray groups. The result for F_4_ demonstrates the most significant difference with *p* = 0.00097 (red: 0.73 ± 0.02, gray group: 0.51 ± 0.01). Although the p values of F_2_ and F_3_ groups were slightly higher than the significance level, the means of the red groups are higher than the those of the gray groups, and the standard deviations of the red groups are much lower than those of the gray groups (F_2_: red 0.87 ± 0.0008, gray 0.73 ± 0.04; F_3_: red 0.91 ± 0.0009, gray 0.84 ± 0.03). In Fig. 10D, the mean iPL of the gray groups are significantly lower than those of the red groups. The results in Fig. 10A and Fig. 10D generally fall in our expectation.

**Fig. 10.**
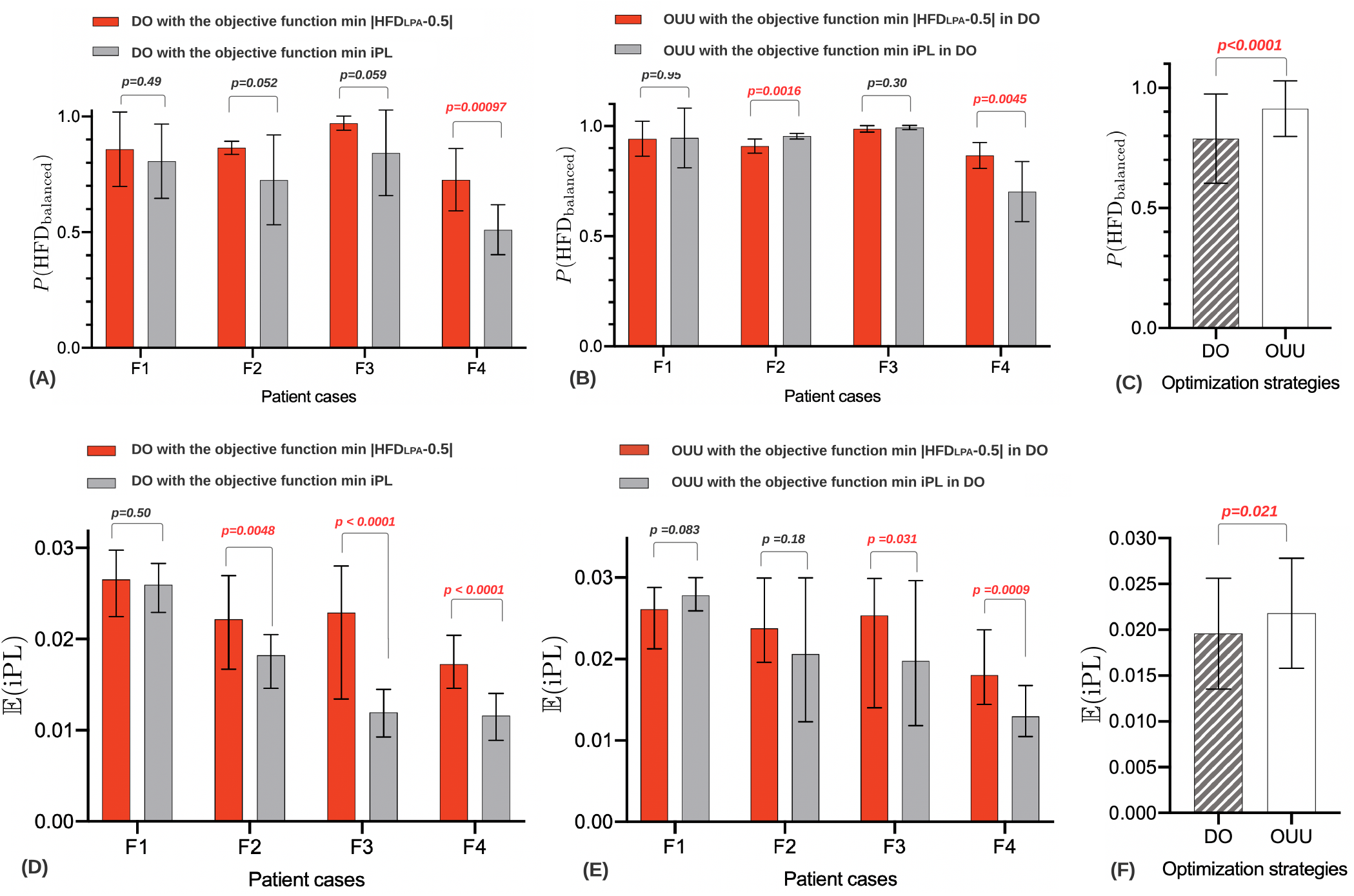
Effect of changing the objective function in DO as minimizing iPL. (A) HFD reliability comparison between DO designs computed from the objective functions min |HFD_LPA_ − 0.5| and min iPL. (B) HFD reliability comparison between OUU designs computed from the two different objective functions in DO. (C) HFD reliability comparison between DO and OUU designs. (D) 𝔼(iPL) comparison between DO designs computed from the two different objective functions in DO. (E) 𝔼(iPL) comparison between OUU designs computed from the two different objective functions in DO. (F) 𝔼(iPL) comparison between DO and OUU designs. A *p*< 0.05 is considered statistically significant.

The OUU performance of different DO objective functions shown in Fig. 10B is mixed for *P*(HFD_balanced_). No significant difference between red and gray groups for the patients F_1_ (*p* = 0.95) and F_3_ (*p* = 0.30). The gray group in F_2_ significantly performs better than the red group (*p* = 0.0016). In contrast, the red group significantly performs better than the gray group in F_4_ (*p* = 0.0045). In Fig. 10E for 𝔼(iPL), the gray groups show significantly better performance than the red groups in F_3_ (*p* = 0.031) and F_4_ (*p* = 0.0009). No significant differences between the red and gray groups are shown in F_1_ (*p* = 0.083) and F_2_ (*p* = 0.18). According to the results, the OUU algorithm does not seem to have a strong preference on the DO objective function.

To demonstrate the effectiveness of the RBRO framework, we combined DO and OUU results of all patients to perform a statistical analysis with 80 conduit designs in each group (n=80). Our results indicate that the conduit designs computed from OUU significantly outperform the designs computed from DO for *P*(HFD_balanced_) with *p* < 0.0001 and 𝔼(iPL) with *p* = 0.021, as shown in Fig. 10C and Fig. 10F.

### D. Comparison of Manually Designed Conduit with DO and OUU designs

We compare the reliability of a manually performed randomized search of conduit design with the best DO and OUU conduit designs for the patient case F_2_. The procedure of manual conduit design involves creating a spectrum of conduit models by using computer-aided design (CAD) software and evaluating them by using CFD software. The conduit with the best performance was selected for manually generating a new group of conduit designs in the next iteration. Three iterations were used to select the best conduit design [8].

To measure *P*(HFD_balanced_), 𝔼(iPL), and 𝔼(%WSS) for the manual conduit design, we generated a parameterized duplicate by minimizing the geometrical difference between the conduits, as shown in Fig. 11A. In Fig. 11B and Fig. 11C, we compare the manually optimized conduit with DO design and OUU design, respectively. As shown in Table II, the manually optimized conduit exhibits lowest reliability in HFD and highest iPL while the OUU design demonstrates 14% improvement on *P*(HFD_balanced_). The OUU design also shows the lowest mean iPL among the three designs.

**Fig. 11.**
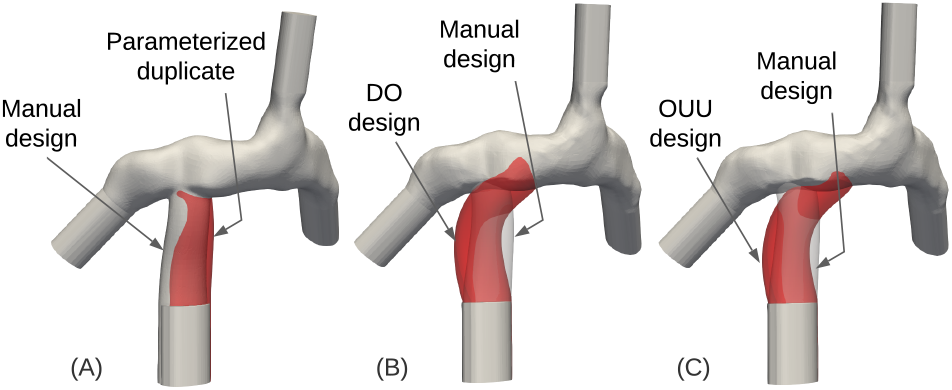
Illustration of Fontan grafts from manual optimization, DO and OUU. (A) Manually optimized graft duplication by minimizing the geometrical difference with the parameterized graft for the patient F_2_. (B) Comparison between the top-ranked DO design and the manually optimized design for F_2_. (C) Comparison between the top-ranked OUU design and the manually optimized design for F_2_.

**TABLE II.**
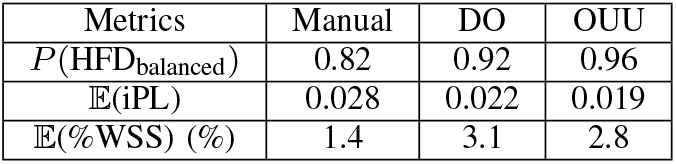
Hemodynamic performance comparison of graft designs from manual optimization, top-ranked DO and top-ranked OUU for F_2_.

| Metrics | Manual | DO | OUU |
| --- | --- | --- | --- |
| $P(\text{HFD}_{\text{balanced}})$ | 0.82 | 0.92 | 0.96 |
| $\mathbb{E}(\text{iPL})$ | 0.028 | 0.022 | 0.019 |
| $\mathbb{E}(\% \text{WSS}) (\%)$ | 1.4 | 3.1 | 2.8 |

## V. Discussion

Virtual surgical planning of Fontan graft implantation involves predicting post-operative outcomes and searching for patient-specific optimal solutions. The uncertainties of cardiovascular system modeling and anastomosis displacements significantly affect the HFD performance of optimal surgical plans that were computed by DO methods [9], [16]. To tackle this challenge, we developed a RBRO framework by actively taking these uncertainties in the Fontan pathway design optimization process and maximizing the probability of keeping HFD within the thresholds for making reliable surgical plans. The study demonstrates that the RBRO framework can significantly improve the reliability of HFD for the optimized Fontan conduits by comparing with the DO framework (*p* < 0.0001).

In Section IV-A, we confirmed that the DO designs of Fontan conduits were unable to reliably provide good hemodynamic performance under the uncertainties. We also noticed that the top-ranked DO designs by using the UQ method demonstrated comparable performance with the top-ranked OUU designs in the four patient cases. It is worth noting that the re-ranking process of DO solutions is inherently an OUU procedure, but with significantly smaller computation cost than a full OUU process. The UQ-based re-ranking of DO solutions could potentially be a more efficient computation strategy for Fontan conduit designs.

The cases of T_1_ and T_2_ in Section IV-B represent Fontan patients with highly unbalanced flow split of PA. We found that for T_1_, which represent an adult patient with *Q*_IVC_/(*Q*_IVC_ + *Q*_SVC_) = 0.72 as shown in Fig. 8, OUU provides little improvement on the reliability of HFD even though the best DO solution of T_1_ was below 0.7 due to the highly unbalanced PA flow split. When we checked on the other adult patient case F_3_, the DO and OUU solutions were close as well, as shown in Fig. 6C. The results imply that the dominant systemic venous flow from IVC may reduce the uncertainty of HFD and leave OUU little room to improve the reliability of DO solutions. In contrast, the IVC and SVC flow rates of the T_2_ case are close (*Q*_IVC_/(*Q*_IVC_ + *Q*_SVC_) = 0.54). The flow competition of IVC and SVC not only results in high power loss [38], but it may also contribute to the low DO design reliability that can be significantly improved by OUU.

The results in Fig. 10 demonstrate that the different objective functions in DO, i.e., min − *P*(HFD_balanced_) and min iPL, were not consistently preferred by OUU in different patient cases. It is most likely because the DO solutions only serve as warm starts of OUU. Depending on the profile of HFD functions related to the design parameters and uncertain parameters, OUU final solution could be adjacent to or far from the initial warm start for searching more reliable conduit designs with a few hundreds iterations.

As we stated in Section II-D, this paper focuses on investigating short-term post-operative OUU results, which means vessel growth, cardiac output changes, increase of *Q*_IVC_/*Q*_SVC_ for pediatric patients were not considered. Due to the lack of data on the uncertain parameters, we assumed that these parameters follow Gaussian distribution according to prior studies [39], [26]. In addition, there is currently a lack of data in the literature characterizing the surgical implantation accuracy. In the current clinical practice, surgeons rely on their experience and using rulers to identify the anastomosis location as prescribed in the virtual surgical planning. A future research study will consider the patient growth in the OUU of Fontan pathway for pediatric patients. The quantification of surgical implantation accuracy is also important to demonstrate the level of conduit implantation error in the current practice.

## VI. Conclusion

Virtual surgical planning and optimization of Fontan grafts could help in improving post-operative hemodynamic performance of the patients. The uncertainties of post-operative blood flow conditions and graft anastomosis displacement may significantly degrade the performance of the prescribed surgical plans that are computed from deterministic patient-specific models. Aiming to address this problem, we developed a RBRO framework for patient-specific Fontan surgical planning. The RBRO framework is capable of automatically computing the patient-specific Fontan conduit with the maximized possibility of keeping all the hemodynamic prameters, including HFD, iPL, and %WSS, in the clinically acceptable range under the presence of uncertain post-operative BC and anastomosis displacements. We tested the proposed RBRO method on four Fontan models that require revision. Compared to the DO conduit designs, the conduit design computed from the proposed method demonstrated significantly improved reliability (up to 79.2%) of HFD, while constraining the mean iPL and %WSS below the threshold. The effectiveness of the proposed method encourages its application to accounting for more challenging conditions, such as the growth of pediatric patients.

## Data Availability

All data produced in the present work are contained in the manuscript

## Acknowledgement

This work was supported by National Institute of Health under grants NHLBI-R01HL143468, R21/R33HD090671, and TEDCO Maryland Innovation Initiative grant 1120-004. The authors acknowledge the supercomputing resource at the Maryland Advanced Research Computing Center (MARCC) (https://www.marcc.jhu.edu/) for providing the computational resources for this study.

